# Genetic architecture of lipoma susceptibility implicates telomere biology

**DOI:** 10.64898/2026.09.08.26362503

**Authors:** David B. Lowry

**Affiliations:** Department of Plant Biology, Michigan State University

## Abstract

Lipomas are common benign adipocytic neoplasms with well-characterized somatic cytogenetic alterations, but the inherited genetic architecture predisposing to their formation remains poorly understood. Recent phenome-wide evidence identified lipoma as strongly associated with genetic predisposition toward longer telomeres. Here, I examined this relationship from the complementary perspective of lipoma genetics. FinnGen R13 genome-wide association and native fine-mapping data resolved nine independent lipoma susceptibility signals across seven genomic regions. Unbiased annotation of these signals implicated telomere-maintenance and genome-stability genes, including telomerase reverse transcriptase (*TERT*), regulator of telomere elongation helicase 1 (*RTEL1*), and STN1 subunit of CST complex (*STN1*), with proximity-expanded mapping additionally encompassing telomerase RNA component (*TERC*). Using 198 leukocyte telomere length (LTL)- associated genetic variants from All of Us, multiplicative-random-effects inverse-variance-weighted Mendelian randomization showed a strong positive association between genetically proxied LTL and lipoma susceptibility (β=0.445, 95% CI 0.355–0.535; P=2.49×10⁻²²). The association replicated using independent UK Biobank LTL effects (β=0.587, 95% CI 0.459–0.715; P=3.09×10⁻¹⁹), remained positive across alternative estimators and sensitivity analyses, persisted after exclusion of 15 variants assigned to canonical telomere/DNA-damage-response genes, and extended across lipomas of the limbs, trunk, and head/face/neck. Regional analyses showed strong evidence for a shared LTL–lipoma association component at *TERC* (PP.H4=0.989), strong but unresolved regional overlap at *TERT* (PP.H4=0.924), and distinct association components at *STN1* (formerly *OBFC1*; PP.H3=0.9995), demonstrating heterogeneous local architectures. The study used a human-supervised agentic research workflow in which ChatGPT contributed to analytical planning, interpretation, quality control, and synthesis, while Codex performed substantial computational execution and reproducibility auditing. Together, these findings connect the independently derived genetic architecture of lipoma susceptibility with a broader trade-off between maintenance of cellular replicative capacity and suppression of neoplastic growth.

## Introduction

Lipomas are among the most common benign mesenchymal neoplasms in humans and consist predominantly of mature adipocytes. Although generally clinically inconsequential, they can occur as solitary or multiple tumors and provide a useful model for understanding the genetic processes that permit clonal growth without malignant transformation. Cytogenetic studies established decades ago that lipomas are genetically heterogeneous neoplasms rather than simple overgrowths of normal adipose tissue. Recurrent somatic alterations include rearrangements of chromosome 12q13–15, abnormalities involving 6p21–23, deletions of chromosome 13q, and several less common cytogenetic classes [1].

The most extensively characterized somatic pathway involves high mobility group AT-hook 2 (*HMGA2*), located at 12q14–15 [2–4]. Chromosomal rearrangements involving this region are common in conventional lipomas and can produce *HMGA2* fusion transcripts or separate the coding region from regulatory sequences in its 3′ untranslated region, resulting in aberrant *HMGA2* expression. The 188-tumor cytogenetic series documented recurrent 12q13–15 abnormalities alongside distinct ring- chromosome, 6p, and chromosome 13 subgroups [1]. Modern sequencing has likewise demonstrated substantial somatic heterogeneity among ordinary lipomas [21]. Thus, considerable progress has been made in defining the genetics of established tumors, while much less is known about inherited variation that determines an individual’s susceptibility to forming lipomas.

Evidence for inherited predisposition comes from familial multiple lipomatosis and the occurrence of multiple lipomas in several genetic syndromes [20,22]. Early cytogenetic work also suggested that multiple and sporadic lipomas might differ biologically: in the Mandahl et al. 188-tumor series, only one of 58 tumors from 18 patients with multiple lipomas had detectable karyotypic changes, leading the authors to propose that a constitutional predisposing mutation might underlie at least some cases [1]. Population-scale genomic resources now provide an opportunity to investigate common inherited susceptibility directly. A focused UK Biobank genome-wide association study (GWAS) identified common-variant associations for lipoma among dermatologic phenotypes, including a signal near low-density lipoprotein receptor-related protein 3 (*LRP3*) [18].

Telomere biology provides a broader framework for considering inherited susceptibility to clonal growth [6]. Progressive telomere shortening limits the proliferative lifespan of somatic cells and can suppress tumor development by triggering senescence or cell death, but the same restriction of proliferative capacity can contribute to declining tissue maintenance and age-related degenerative disease [23,32,33]. This tension has been described as an evolutionary trade-off between tumor suppression and maintenance of regenerative capacity. Consistent with that model, Mendelian-randomization studies have found that genetically longer telomeres are associated with increased risk of several cancers while reducing risk for some non-neoplastic diseases [23–25]. Recent work further emphasizes telomere attrition as a broadly acting tumor-suppressive barrier that limits the outgrowth of cells with oncogenic potential [33].

Particularly intriguing evidence that this trade-off may extend to benign neoplasia came from Allaire et al. [19]. In a phenome-wide analysis of 113,861 All of Us participants, genetic predisposition toward longer telomeres was associated with a broad spectrum of benign and malignant tumors, with lipoma showing the strongest association with the TOPMed- derived telomere polygenic score (P=4.2×10⁻¹⁵). Allaire et al. proposed a “long-telomeropathy” model in which genetically increased telomere length contributes to tumor susceptibility, potentially by extending cellular replicative potential. Lipoma is especially informative in this context because it represents clonal neoplastic growth without the invasive and metastatic properties that define cancer, raising the possibility that benign tumors occupy part of the same biological continuum between preservation of replicative capacity and suppression of abnormal clonal expansion.

Here, I examined the telomere–lipoma connection from the complementary perspective of lipoma genetics itself. I used FinnGen R13 genome-wide association and fine-mapping data to characterize the germline architecture of benign lipoma and asked whether independently identified susceptibility loci converge on telomere-related biology. Motivated by that convergence, I tested the relationship between leukocyte telomere length (LTL) genetics and lipoma susceptibility using exact All of Us LTL summary statistics with FinnGen as an independent outcome, replicated the analysis using UK Biobank LTL effects, examined anatomical classes of lipoma, and investigated local genetic architecture at prominent telomere-associated loci. Together, these analyses test whether a common benign neoplasm can illuminate the broader trade-off between cellular replicative potential and protection from neoplastic growth.

This study used a human-supervised agentic research workflow involving ChatGPT and Codex; I retained responsibility for scientific decisions, interpretation, and final claims. Details are provided in Methods.

## Methods

### Study design and genetic data sources

I designed the analysis to approach the telomere–lipoma relationship from two complementary directions. First, I characterized the inherited genetic architecture of lipoma without using telomere-length results to select loci or genes. Second, after telomere-maintenance genes emerged from that analysis, I tested whether genetic variants associated with LTL were also associated with lipoma susceptibility. The primary lipoma outcome was FinnGen release 13 broad benign lipomatous disease (endpoint `CD2_BENIGN_LIPOMATOUS`), comprising 13,526 cases and 486,660 controls. FinnGen association statistics were analyzed on the GRCh38 coordinate system and retained on the reported log-odds scale [26]. I also used FinnGen phenotypes for lipomas of the limbs, trunk, and head/face/neck to ask whether the LTL association generalized across anatomical sites; phenotype-specific case/control counts were not retained in the analysis summaries and are therefore not reported here.

To define genetic proxies for LTL, I started with 210 independently associated European variants reported in Supplementary Table 7 of Nakao et al. [7]. Exact All of Us EUR LTL GWAS summary statistics were obtained from the public Nakao et al. Zenodo release (record 16689623; DOI `10.5281/zenodo.16689623`). The local copy was verified against the released GRCh38 file by MD5 checksum (`75ff7d23bc0dcf001b59cdcdf6c4fcda`). I did not access individual-level All of Us participant data. For independent replication, I used the corresponding UK Biobank EUR LTL summary statistics from the same public release. Because the All of Us and UK Biobank effect estimates are not documented on an identical standardized scale, I report MR effects on the study-specific LTL scales rather than per standard deviation.

### Fine-mapping and functional interpretation of lipoma susceptibility loci

To distinguish independent lipoma susceptibility signals from clusters of correlated genome-wide-significant variants, I used FinnGen’s native SuSiE and FINEMAP fine-mapping outputs [12,13,27]. FinnGen defines fine-mapping regions around genome-wide-significant lead variants, calculates in-sample dosage linkage disequilibrium (LD) with LDstore2, and applies both FINEMAP and SuSiE while allowing multiple causal variants per region [27]. I treated the native SuSiE credible sets as the units of independent lipoma association. For each signal, I recorded the 95% credible-set size, lead-variant posterior inclusion probability (PIP), lead association P value, and average and minimum pairwise r² within the credible set. Nine signals across seven genomic regions were identified. The accessible project records do not establish the exact R13 production container or LD-panel implementation beyond the publicly documented FinnGen pipeline, so those details are not inferred here.

To ask what biology was represented by the lipoma susceptibility loci without allowing the subsequent LTL analyses to influence gene selection, I annotated all 326 variants in the nine 95% SuSiE credible sets before interpreting the telomere-length analyses. I used two prespecified coordinate-based mappings on GRCh38. **Direct-overlap mapping** assigned a gene only when a credible-set variant physically overlapped its Ensembl gene interval. **Proximity-expanded mapping** additionally included genes located within 100 kb of a credible-set variant. Gene intervals were obtained with the Ensembl REST application programming interface using GRCh38 release 116 [29,30]. I then evaluated Gene Ontology Biological Process, Reactome, KEGG, and WikiPathways annotations using g:Profiler [31]. Because the effective independent sample consisted of nine loci rather than the individual genes within them, I treated pathway P values as descriptive evidence of biological convergence rather than as definitive independent enrichment tests. Complete coding, regulatory, expression quantitative trait locus (eQTL), and splicing quantitative trait locus (sQTL) annotation was not available.

### Mendelian-randomization analysis of LTL and lipoma susceptibility

To test whether genetic predisposition toward longer LTL was associated with lipoma susceptibility, I matched the 210 published LTL-associated variants to the exact All of Us EUR summary statistics by genomic position and alleles. Effect estimates were oriented to the published LTL-increasing allele, complementing allele frequency when required by strand orientation. Two hundred variants were present in the exact All of Us data, and 198 of these could also be matched unambiguously to the FinnGen lipoma GWAS. Outcome effects were aligned to the same effect allele. These 198 LTL-associated variants constituted the primary MR set.

I estimated the association using inverse-variance-weighted (IVW) Mendelian randomization (MR), reporting both fixed-effect and multiplicative-random-effects (MRE) models. The IVW point estimate is the same under these implementations, but the MRE model inflates uncertainty when variant-specific ratio estimates are heterogeneous. Because Cochran’s Q indicated substantial heterogeneity, I used the MRE confidence interval and P value as the principal inference. To evaluate whether the result depended strongly on IVW assumptions, I also used the weighted-median estimator and Mendelian randomization-Egger (MR-Egger) regression [8,9,11]. Cochran’s Q quantified heterogeneity, and the MR-Egger intercept tested for average directional horizontal pleiotropy [10]. A nonsignificant intercept was not interpreted as evidence that pleiotropy was absent.

To assess whether a small number of variants dominated the association, I repeated the IVW analysis after omitting each of the 198 LTL-associated variants in turn. I also used RadialMR as a complementary diagnostic for variants whose effects were poorly fit by the overall IVW relationship [16]. Radial outliers were treated as diagnostic flags rather than automatically excluded variants. MR-PRESSO was explored as an additional pleiotropy/outlier analysis, but higher-precision simulation runs were computationally impractical; its lower-precision results were therefore not used to select variants for the primary analysis [10].

To test whether the association was simply a consequence of the well-known telomere-maintenance loci that initially drew attention to the phenotype, I performed a prespecified exclusion analysis. Fifteen of the 198 LTL-associated variants were assigned to genes in a canonical telomere/DNA-damage-response (DDR) set (Supplementary Table 4). I repeated the MR analysis after removing those 15 variants, leaving 183 variants. The classification was defined independently of the lipoma effect estimates.

Finally, I used Steiger directionality analysis to ask whether the aggregate genetic association was more consistent with LTL influencing lipoma than the reverse [28]. The comparison used variance explained in LTL and an approximation of variance explained in the binary FinnGen lipoma outcome based on the observed case fraction. Because that approximation does not provide a population liability-scale estimate, I treated the Steiger result as supportive evidence about direction rather than proof of causality.

### Replication and anatomical analyses

To determine whether the LTL–lipoma association depended on the All of Us estimates of genetic effects on telomere length, I repeated the analysis using independently estimated LTL effects from UK Biobank. I matched the same published LTL-associated variants to the UK Biobank EUR summary statistics and retained the same 198 variants for which compatible FinnGen lipoma data were available. Keeping the variant set fixed ensured that differences between the All of Us and UK Biobank analyses reflected independently estimated LTL effects rather than selection of different genetic variants. I compared the direction and correlation of variant-specific LTL effects between the two exposure datasets and repeated the IVW-MRE, weighted-median, and MR-Egger analyses.

To determine whether the association was specific to lipomas at one anatomical site, I applied the unchanged 198-variant framework to the FinnGen limb, trunk, and head/face/neck lipoma phenotypes. For each site and exposure dataset, I estimated fixed-effect and MRE IVW associations and calculated Cochran’s Q. I compared the site-specific estimates descriptively; I did not perform a formal between-site meta-regression.

### Locus-level colocalization and linkage analyses

A genome-wide MR association can arise even when individual loci relate to exposure and outcome in different ways. At a region associated with both LTL and lipoma, the two traits could be influenced by the same causal variant (local pleiotropy), by distinct causal variants that are correlated through LD (linkage), or by more complex multi-signal architecture. To distinguish among these possibilities, I examined three prominent telomere-associated lipoma regions centered on telomerase RNA component (*TERC*), telomerase reverse transcriptase (*TERT*), and STN1 subunit of CST complex (*STN1*; formerly *OBFC1*).

I used existing regional association statistics for FinnGen lipoma and All of Us LTL in classical coloc analyses [14]. The coloc framework compares posterior support for alternative configurations, including H3, in which both traits are associated but with distinct causal variants, and H4, in which the traits share a causal variant. I interpreted these probabilities as evidence about regional architecture rather than definitive causal-variant assignment because the analyses were not uniformly multi-signal SuSiE-coloc analyses. At *TERT*, where the leading lipoma and shared-colocalization variants differed, I additionally examined pairwise Finnish LD. Moderate LD between the candidate variants was considered insufficient to treat them as the same signal. Exact software and panel settings for this pairwise LD query were not retained, so the LD result is used only as supporting regional evidence.

### Exploratory comparison with uterine leiomyoma

To ask whether lipoma susceptibility might form part of a broader inherited architecture of benign mesenchymal tumors, I performed an exploratory comparison with FinnGen R13 uterine leiomyoma (42,816 cases and 239,093 controls). I compared the nine lipoma SuSiE regions with leiomyoma regional associations and assessed whether regional overlap exceeded that expected under a coarse interval-permutation procedure. This analysis was intentionally treated as exploratory because native leiomyoma signal-level SuSiE/FINEMAP membership and corresponding Finnish LD resources were not available locally. I therefore did not infer shared causal variants or genes from regional coincidence alone.

### Statistical software, reproducibility, and agentic workflow

Core MR analyses were performed in R 4.5.2 using TwoSampleMR 0.7.9, MendelianRandomization 0.10.0, MR-PRESSO 1.0, and RadialMR; colocalization used coloc 5.2.3. Data harmonization, annotation, quality-control auditing, and figure generation used R and Python 3.14.3. The exact historical RadialMR and matplotlib versions were not retained. Analysis scripts, detailed software and data provenance, derived data underlying the figures and tables, and reproducibility audits are available in the accompanying public repository.

This project used ChatGPT and Codex as active components of a human-supervised agentic research workflow rather than solely for language editing. ChatGPT contributed iteratively to analytical planning, selection and evaluation of statistical approaches and public datasets, interpretation of intermediate results, quality-control review, follow-up prioritization, scientific synthesis, and manuscript development. Codex performed substantial computational execution within the project workspace, including data inspection, scripting, harmonization, auditing, table and figure generation, and reproducibility checks. I retained responsibility for the scientific questions, analysis decisions, interpretation, and final claims. The ChatGPT model used during the current manuscript-development phase was GPT-5.6 Sol. The underlying model identifier used during historical Codex computational sessions was not retained and therefore cannot be reported retrospectively. Additional software and agentic-workflow provenance is provided in the accompanying repository.

To reduce the risk of propagating errors through the agentic workflow, I checked key outputs against source data and scripts, preserved superseded outputs during auditing, and investigated discrepancies rather than silently overwriting them. This process identified and corrected, among other issues, an MR effect discrepancy caused by rounded versus exact All of Us exposure effects and an incorrect count in the telomere/DDR exclusion sensitivity analysis. Negative or ambiguous results, including the *STN1* colocalization and leiomyoma enrichment analyses, were retained. ChatGPT and Codex are research tools and are not authors of this work.

### Ethics statement

This study used publicly available aggregate genetic summary statistics and did not access individual-level participant data.

## Results

### FinnGen identifies a heterogeneous genetic architecture of lipoma susceptibility

I characterized germline susceptibility to lipoma using the FinnGen R13 broad benign lipomatous tumor phenotype, comprising 13,526 cases and 486,660 controls. Genome-wide association signals occurred across multiple chromosomes, and native FinnGen SuSiE fine-mapping resolved nine independent signals across seven genomic regions (Fig. 1; Table 2). The nine lipoma signals differed markedly in credible-set size and fine-mapping resolution.

**Figure 1.**
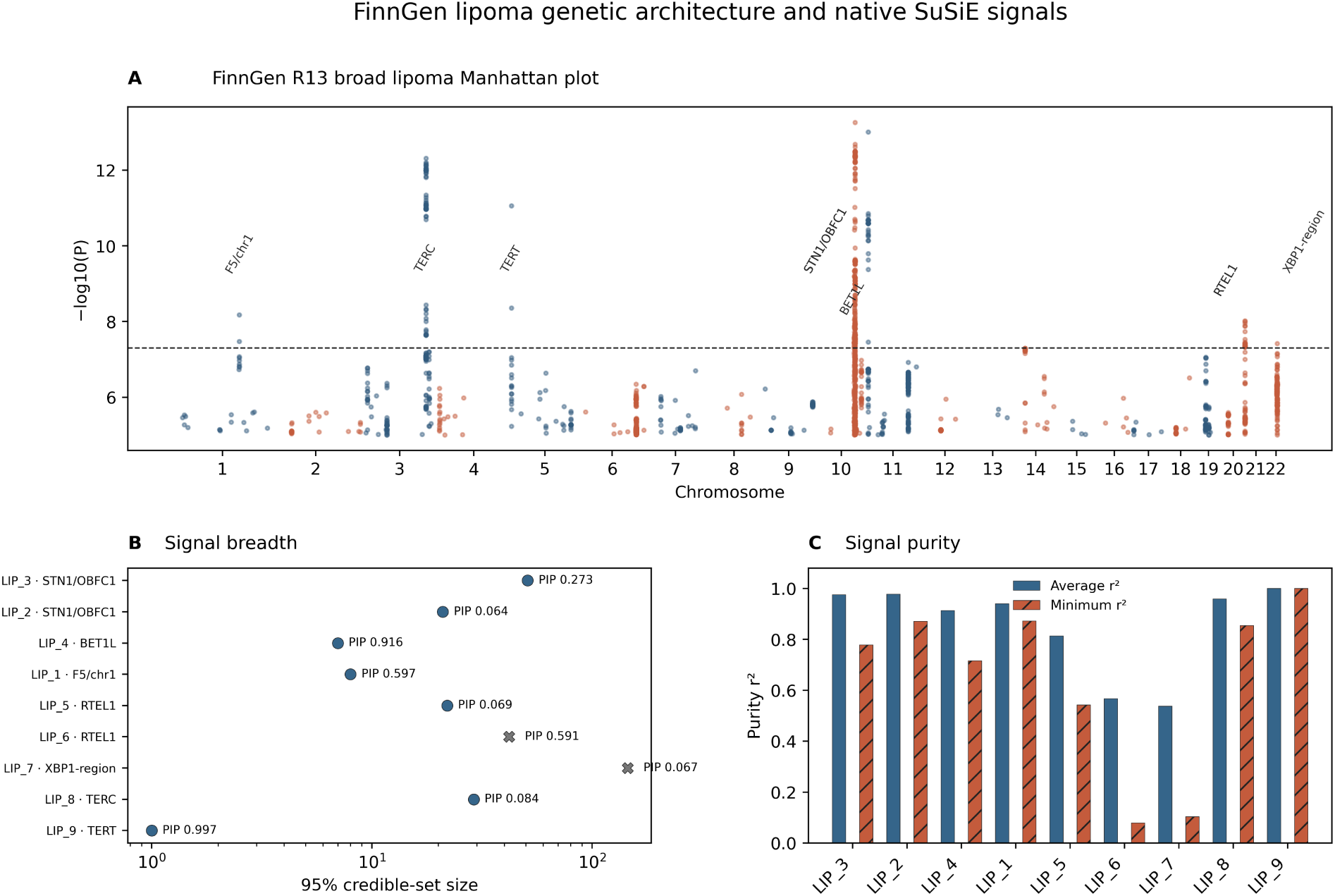
FinnGen R13 broad lipoma association and native SuSiE fine-mapping results. (A) Conventional cumulative- position Manhattan plot for chromosomes 1–22. The x-axis is cumulative genomic position, with chromosome numbers shown at chromosome centers; the y-axis is −log10(P). Alternating blue and orange points distinguish adjacent chromosomes and do not encode effect direction or probability. The dashed line marks P=5×10⁻8. Labels identify seven fine-mapped regions and are conservative interpretive descriptors, not causal-gene assignments. (B) The nine independent SuSiE signals are shown by 95% credible-set size on a logarithmic x-axis; text gives lead-variant PIP. Circles denote signals not flagged for low purity and X markers denote low-purity signals. (C) Bars show mean pairwise r² (solid blue) and minimum pairwise r² (hatched orange) within each credible set. PIP, posterior inclusion probability; r², squared pairwise dosage correlation; SuSiE, Sum of Single Effects. The lead variant is not necessarily causal. Nine signals and 326 credible-set variants are represented.

**Table 1.**
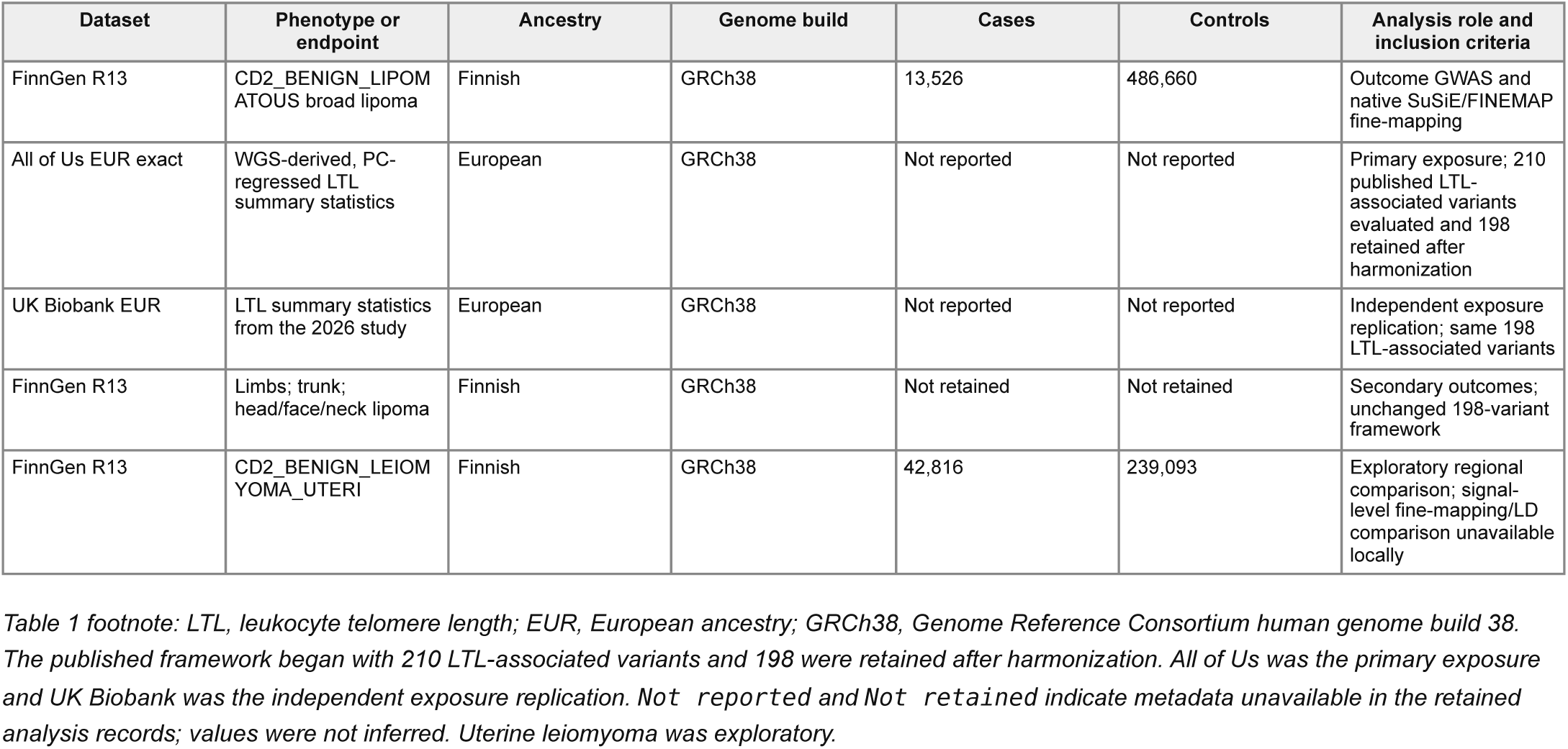
Genetic data resources used to characterize lipoma susceptibility and leukocyte telomere length.

**Table 2.**
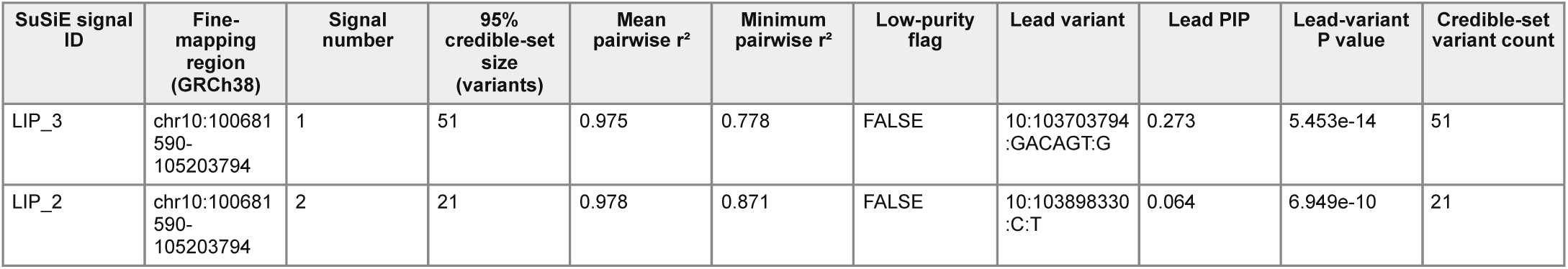
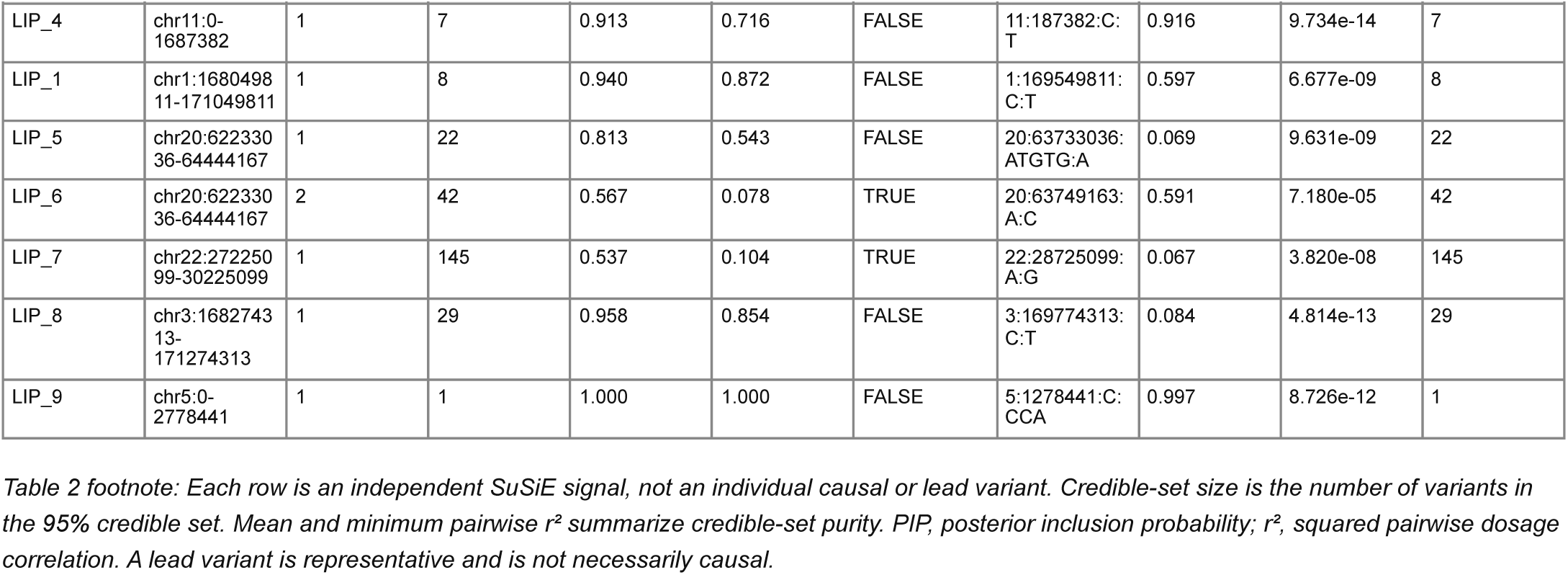
Nine independent FinnGen R13 SuSiE signals define the germline genetic architecture of lipoma susceptibility.

The most precisely resolved association occurred on chromosome 5, where the 95% credible set consisted of a single variant, 5:1278441:C:CCA, with posterior inclusion probability (PIP) of 0.997. A compact chromosome 11 signal contained seven variants and was dominated by 11:187382:C:T (PIP=0.916), while the chromosome 1 signal contained eight variants with a lead-variant PIP of 0.597. In contrast, the chromosome 22 signal contained 145 variants and had low purity (minimum r²=0.104), indicating poor causal resolution.

Several regions showed evidence of more complex local architecture. Two independent signals were identified within the chromosome 10 region, with 51- and 21-variant credible sets, respectively. Two signals were likewise resolved near *RTEL1* on chromosome 20, including one with particularly low purity (minimum r²=0.078). The chromosome 3 signal near *TERC* contained 29 variants with high credible-set purity (minimum r²=0.854), but posterior probability was distributed among correlated variants rather than concentrated on a single candidate (maximum PIP=0.084). Thus, lipoma susceptibility is characterized by multiple independent loci ranging from highly resolved single-variant signals to broad and multi-signal regions in which causal-variant assignment remains uncertain.

### Independent annotation of lipoma susceptibility loci converges on telomere-maintenance and DNA-repair biology

To investigate the biology represented by the lipoma susceptibility loci without using subsequent LTL analyses to select candidate genes, I performed a prespecified coordinate-based annotation of all 326 variants contained within the nine 95% SuSiE credible sets. Gene-mapping rules were defined before pathway inspection and did not incorporate LTL-associated variants, the previously curated telomere/DNA-damage-response (DDR) gene set, or the leiomyoma results. Direct-overlap mapping assigned genes only when a credible-set variant physically overlapped the annotated gene interval, whereas proximity-expanded mapping additionally included genes within 100 kb of a credible-set variant.

Direct-overlap mapping identified 30 genes and independently recovered several genes with established roles in telomere maintenance or genome stability, including *TERT*, *RTEL1*, and *STN1* [15,17]. Expanded mapping additionally encompassed the *TERC* region. Broad gene-set analysis consequently identified telomere extension, telomere maintenance, telomere organization, replicative senescence, and DNA metabolic processes among the strongest annotations. DNA-repair annotations additionally included checkpoint kinase 2 (*CHEK2*) and SWI5-dependent homologous recombination repair protein 1 (*SFR1*). Because these mapped genes derive from only nine independent susceptibility signals and multiple genes within loci are non-independent, these results are interpreted as qualitative biological convergence rather than definitive statistical evidence for pathway enrichment.

Other biological annotations were less stable across mapping definitions. Extracellular-matrix and vascular-associated terms appeared with direct-overlap mapping, while proximity-expanded mapping additionally identified immune/host-response and cell-cycle-related terms. X-box binding protein 1 (*XBP1*), located within the poorly resolved chromosome 22 region, was the clearest adipocyte-related candidate, but adipocyte-differentiation and mesenchyme-development annotations provided little evidence for a general adipogenic pathway. The independent emergence of multiple telomere-maintenance loci from the lipoma fine-mapping results motivated a direct test of whether inherited genetic variation influencing LTL is associated with lipoma susceptibility.

### Genetically proxied longer leukocyte telomere length is associated with increased lipoma susceptibility

I evaluated 210 previously reported European LTL-associated variants, of which 198 could be harmonized to exact All of Us LTL summary statistics and the FinnGen R13 lipoma GWAS. Genetically proxied longer LTL was strongly associated with increased lipoma susceptibility (Fig. 2; Table 3). Because variant-specific estimates exhibited substantial heterogeneity, I used multiplicative-random-effects inverse-variance weighting (IVW-MRE) as the principal inferential model. The All of Us analysis yielded β=0.445 (95% CI 0.355-0.535; P=2.49×10⁻22). Fixed-effect IVW produced the same point estimate with a narrower confidence interval (SE=0.0325; P=1.58×10⁻42).

**Figure 2.**
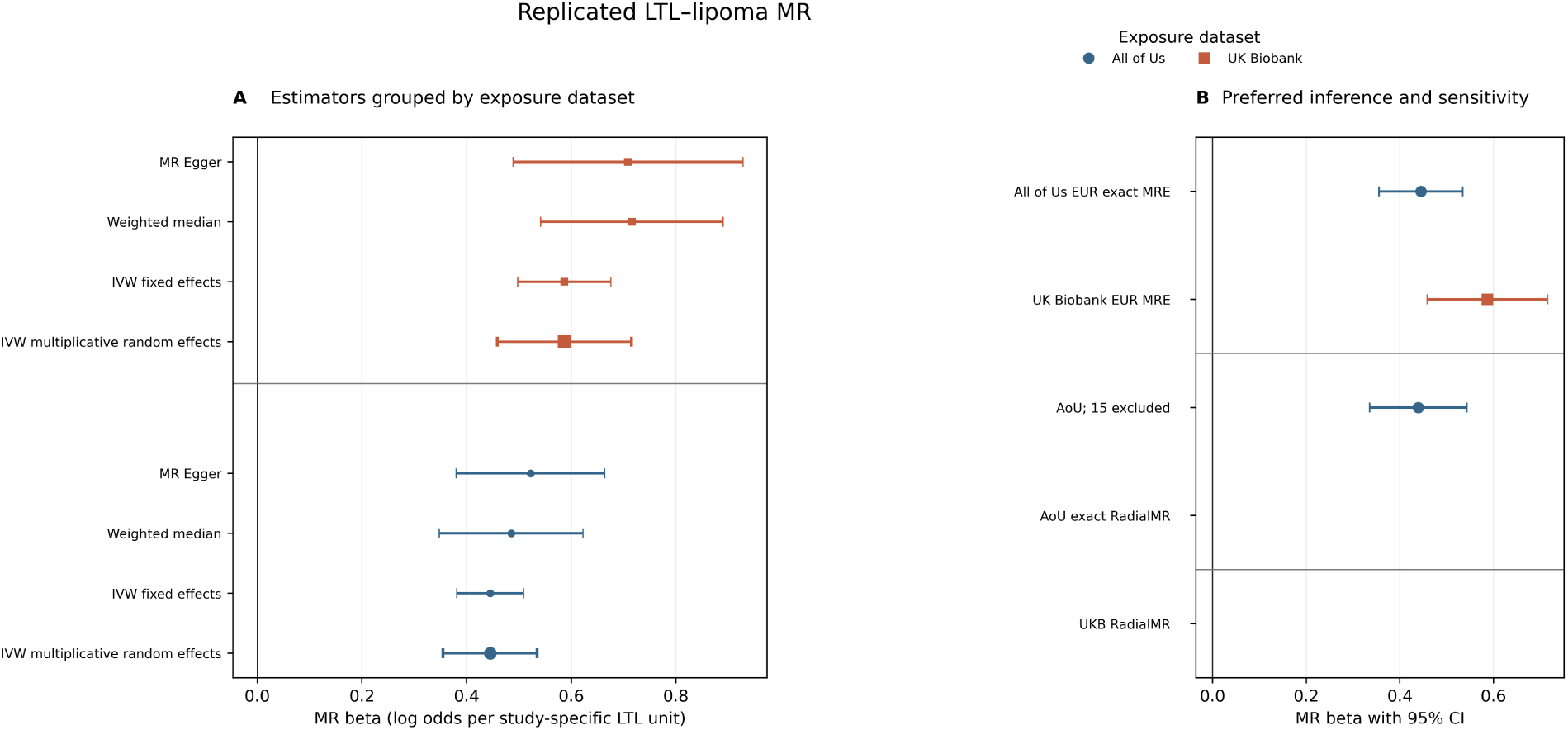
Mendelian-randomization estimates for the association between genetically proxied leukocyte telomere length (LTL) and FinnGen broad lipoma susceptibility. (A) Estimates and 95% confidence intervals are grouped by exposure dataset: blue circles, exact All of Us EUR; orange squares, independent UK Biobank EUR. Rows show IVW multiplicative random effects (MRE), fixed-effect IVW, weighted median, and MR-Egger. MRE is emphasized by marker size because heterogeneity is substantial; this graphical emphasis does not itself establish statistical priority. (B) All of Us MRE, All of Us after excluding 15 canonical telomere/DDR LTL-associated variants, UK Biobank MRE, and RadialMR iterative estimates. Error bars are 95% confidence intervals; sensitivity intervals use the reported beta and SE. The x-axis is MR beta per unit increase in genetically proxied LTL on the relevant study-specific LTL scale. The vertical line is the null. RadialMR flags are diagnostic and no variants were automatically removed. Q, Cochran heterogeneity statistic; DDR, DNA-damage response; IVW, inverse-variance weighted; MR, Mendelian randomization; MRE, multiplicative random effects.

**Table 3.**
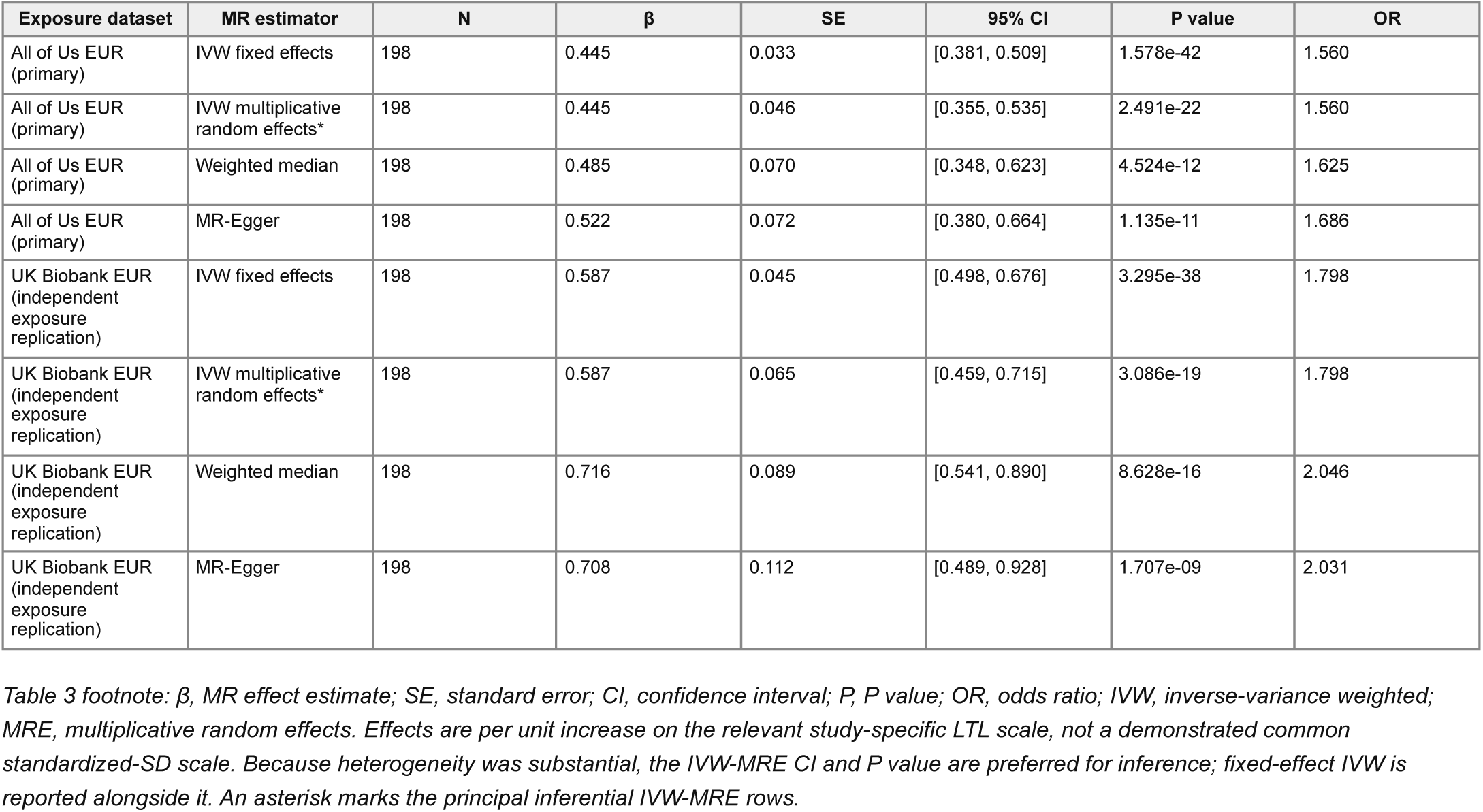
Mendelian-randomization estimates for the association between genetically proxied LTL and lipoma susceptibility in the primary All of Us analysis and UK Biobank replication.

footnote: $\beta$ , MR effect estimate; SE, standard error; CI, confidence interval; P, P value; OR, odds ratio; IVW, inverse-variance weighted; MRE, multiplicative random effects.
| Exposure dataset | MR estimator | N | $\beta$ | SE | 95% CI | P value | OR |
| --- | --- | --- | --- | --- | --- | --- | --- |
| All of Us EUR (primary) | IVW fixed effects | 198 | 0.445 | 0.033 | [0.381, 0.509] | 1.578e-42 | 1.560 |
| All of Us EUR (primary) | IVW multiplicative random effects* | 198 | 0.445 | 0.046 | [0.355, 0.535] | 2.491e-22 | 1.560 |
| All of Us EUR (primary) | Weighted median | 198 | 0.485 | 0.070 | [0.348, 0.623] | 4.524e-12 | 1.625 |
| All of Us EUR (primary) | MR-Egger | 198 | 0.522 | 0.072 | [0.380, 0.664] | 1.135e-11 | 1.686 |
| UK Biobank EUR (independent exposure replication) | IVW fixed effects | 198 | 0.587 | 0.045 | [0.498, 0.676] | 3.295e-38 | 1.798 |
| UK Biobank EUR (independent exposure replication) | IVW multiplicative random effects* | 198 | 0.587 | 0.065 | [0.459, 0.715] | 3.086e-19 | 1.798 |
| UK Biobank EUR (independent exposure replication) | Weighted median | 198 | 0.716 | 0.089 | [0.541, 0.890] | 8.628e-16 | 2.046 |
| UK Biobank EUR (independent exposure replication) | MR-Egger | 198 | 0.708 | 0.112 | [0.489, 0.928] | 1.707e-09 | 2.031 |

Alternative estimators supported the same direction of association. The weighted-median estimate was β=0.485 (95% CI 0.348-0.623; P=4.52×10⁻12), and MR-Egger yielded β=0.522 (95% CI 0.380-0.664; P=1.14×10⁻11). Thus, the positive association was not specific to the IVW estimator. Because the All of Us and UK Biobank LTL effects are expressed on study-specific scales rather than a demonstrably common standardized scale, effect magnitudes were interpreted within each exposure dataset rather than as effects per standard deviation of LTL.

### The LTL–lipoma association replicates using independent UK Biobank exposure genetics

I next tested the association using LTL effects independently estimated in UK Biobank. All 198 LTL-associated variants used in the primary analysis were retained, and their LTL effects were directionally concordant between All of Us and UK Biobank. SNP-specific exposure effects were strongly correlated between datasets (Pearson r=0.957; 198/198 concordant signs), although their magnitudes differed systematically, consistent with differences in study-specific LTL scaling (Fig. 3A).

**Figure 3.**
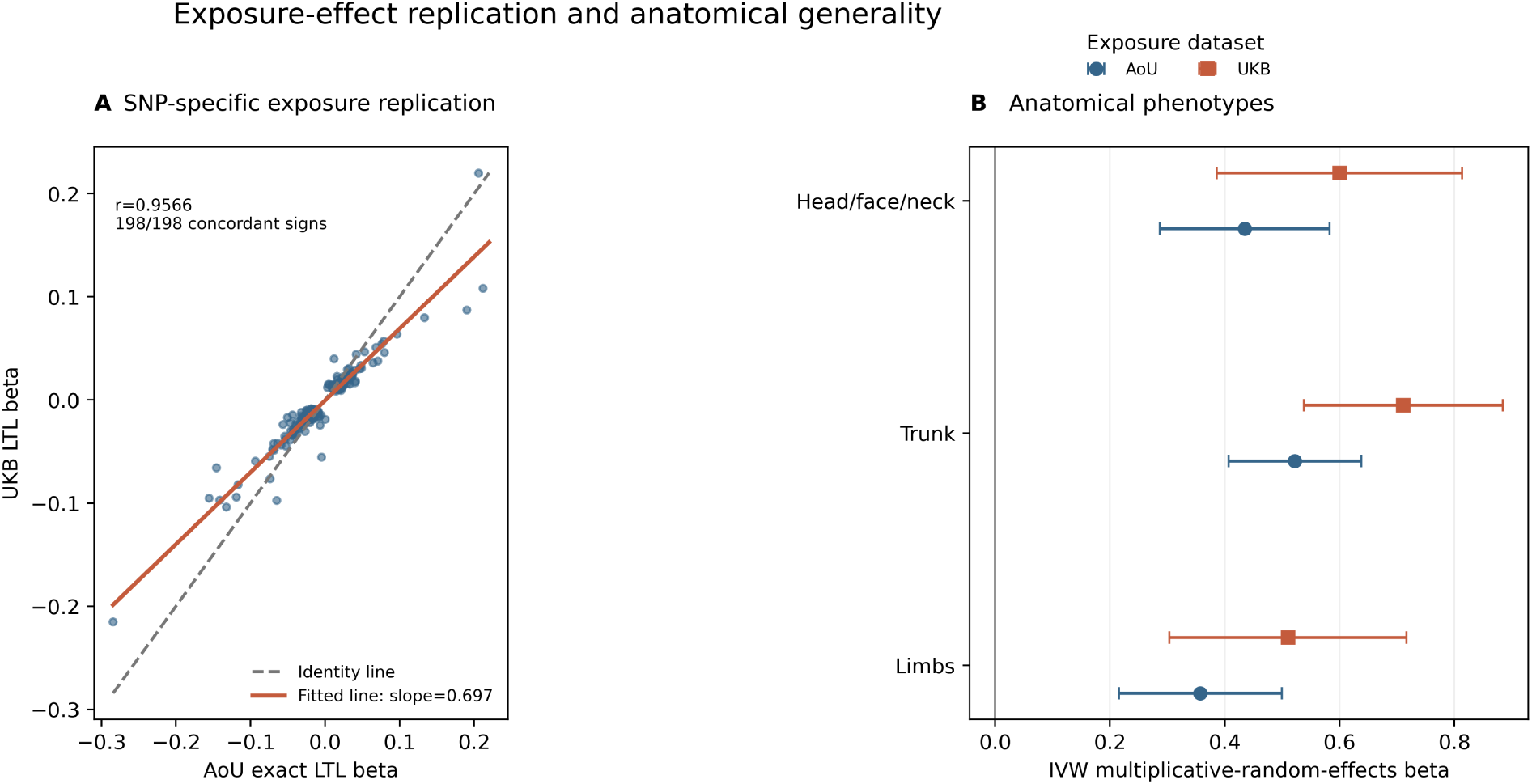
(A) SNP-specific comparison of exact All of Us and independent UK Biobank LTL effects for 198 LTL-associated variants. Blue points are individual variants; the grey dashed line is the identity line and the solid orange line is the fitted UKB-on-AoU regression (slope approximately 0.697). Axes are study-specific LTL beta scales; the annotation gives Pearson r and sign concordance. (B) Anatomical-site IVW-MRE estimates using the unchanged 198-variant set. Rows are Limbs, Trunk, and Head/face/neck; blue circles denote All of Us and orange squares denote UK Biobank. Error bars are 95% confidence intervals. The x-axis is IVW multiplicative-random-effects beta on the relevant study-specific LTL scale; the vertical line is the null. Cochran Q statistics are retained in Table 4 and are not visibly annotated in this panel. LTL, leukocyte telomere length; IVW-MRE, inverse-variance-weighted multiplicative-random-effects model; Q, Cochran heterogeneity statistic.

**Table 4.** Anatomical-site Mendelian-randomization estimates for the association between genetically proxied LTL and lipoma susceptibility.

footnote: All estimates use the same 198 LTL-associated variants.
| Anatomical phenotype | Exposure dataset | MR estimator | N | $\beta$ | SE | 95% CI | P value | Cochran Q | Q P value |
| --- | --- | --- | --- | --- | --- | --- | --- | --- | --- |
| Limbs | All of Us EUR (primary) | IVW multiplicative random effects | 198 | 0.358 | 0.072 | 0.216–0.499 | 7.925e-07 | 223.429 | 9.520e-02 |
| Limbs | UK Biobank EUR (independent exposure replication) | IVW multiplicative random effects | 198 | 0.510 | 0.105 | 0.303–0.717 | 1.324e-06 | 246.862 | 9.145e-03 |
| Trunk | All of Us EUR (primary) | IVW multiplicative random effects | 198 | 0.522 | 0.059 | 0.407–0.638 | 8.278e-19 | 262.600 | 1.227e-03 |
| Trunk | UK Biobank EUR (independent exposure replication) | IVW multiplicative random effects | 198 | 0.711 | 0.088 | 0.538–0.884 | 9.041e-16 | 309.163 | 5.644e-07 |
| Head/face/neck | All of Us EUR (primary) | IVW multiplicative random effects | 198 | 0.435 | 0.076 | 0.287–0.583 | 8.514e-09 | 186.754 | 6.885e-01 |
| Head/face/neck | UK Biobank EUR (independent exposure replication) | IVW multiplicative random effects | 198 | 0.600 | 0.109 | 0.386–0.813 | 3.606e-08 | 212.734 | 2.102e-01 |

Using UK Biobank LTL effects, IVW-MRE again showed a strong positive association with lipoma susceptibility (β=0.587, 95% CI 0.459-0.715; P=3.09×10⁻19). Weighted-median (β=0.716, 95% CI 0.541-0.890; P=8.63×10⁻16) and MR-Egger (β=0.708, 95% CI 0.489-0.928; P=1.71×10⁻9) estimates were likewise positive. The independent exposure replication therefore reproduced the direction and strong statistical support of the All of Us analysis despite differences in LTL effect scaling between cohorts.

### The LTL–lipoma association is robust to influential loci and alternative MR assumptions

Variant-specific estimates showed substantial heterogeneity in the primary All of Us analysis (Cochran’s Q=389.6, 197 df, P=9.04×10⁻15). The MR-Egger intercept did not differ significantly from zero (intercept=-0.00337, SE=0.00245, P=0.170), providing no evidence for directional horizontal pleiotropy by this test, although this result does not exclude pleiotropic effects more generally. Steiger analysis supported the hypothesized LTL-to-lipoma direction, with greater aggregate variance explained in the exposure than in the outcome (R²=0.0725 versus 0.00676), subject to assumptions required to approximate variance explained for the binary lipoma phenotype.

I next asked whether the association was disproportionately driven by variants assigned to canonical telomere and DDR genes. Removal of the 15 prespecified LTL-associated variants in this gene set (Supplementary Table 4) left 183 variants. The resulting IVW-MRE estimate was essentially unchanged (β=0.439, SE=0.0530; P=1.06×10⁻16) relative to the primary β=0.445 estimate. Thus, the genome-wide relationship between LTL genetics and lipoma susceptibility was not dependent on the canonical telomere/DDR loci themselves.

Leave-one-out analyses similarly showed that no single LTL-associated variant accounted for the association. Sequential omission of each of the 198 variants yielded IVW estimates ranging from β=0.413 to 0.463, all in the same direction as the complete analysis. After exclusion of the canonical telomere/DDR set, leave-one-out estimates remained positive (β=0.391- 0.459). RadialMR provided an additional heterogeneity-robust assessment and produced an iterative All of Us estimate closely matching the primary result (β=0.446, SE=0.0458), despite identifying multiple variants with outlying behavior. I therefore retained the complete prespecified variant set for primary inference rather than selecting variants according to their association with the lipoma outcome. Exploratory MR-PRESSO results were not used for variant selection because higher-precision simulation runs could not be completed.

### The association extends across anatomical classes of lipoma

I next asked whether the association was restricted to lipomas arising at a particular anatomical site. Using the unchanged 198-variant framework, I analyzed FinnGen phenotypes for lipomas of the limbs, trunk, and head/face/neck (Fig. 3B; Table 4). IVW-MRE estimates were positive for every anatomical class using both LTL exposure datasets.

Using All of Us LTL effects, estimates were β=0.358 for limb lipomas (95% CI 0.216-0.499; P=7.92×10⁻7), β=0.522 for trunk lipomas (95% CI 0.407-0.638; P=8.28×10⁻19), and β=0.435 for head/face/neck lipomas (95% CI 0.287-0.583; P=8.51×10⁻9). UK Biobank exposure effects produced the same pattern: β=0.510 for limb lipomas (95% CI 0.303-0.717; P=1.32×10⁻6), β=0.711 for trunk lipomas (95% CI 0.538-0.884; P=9.04×10⁻16), and β=0.600 for head/face/neck lipomas (95% CI 0.386- 0.813; P=3.61×10⁻8).

Heterogeneity varied among anatomical phenotypes and was strongest for trunk lipomas, whereas it was not significant for the head/face/neck phenotype in either exposure dataset. These results indicate that the LTL association is not confined to one anatomical class of lipoma, although its magnitude and heterogeneity vary among sites.

### Telomere-associated lipoma loci show heterogeneous local genetic architectures

Finally, I investigated whether prominent telomere-related regions showed evidence for shared local genetic architecture between LTL and lipoma susceptibility. The results differed markedly among loci, demonstrating that the genome-wide association between genetically proxied LTL and lipoma does not imply a uniform relationship at individual telomere- maintenance loci (Fig. 4; Table 5).

**Figure 4.**
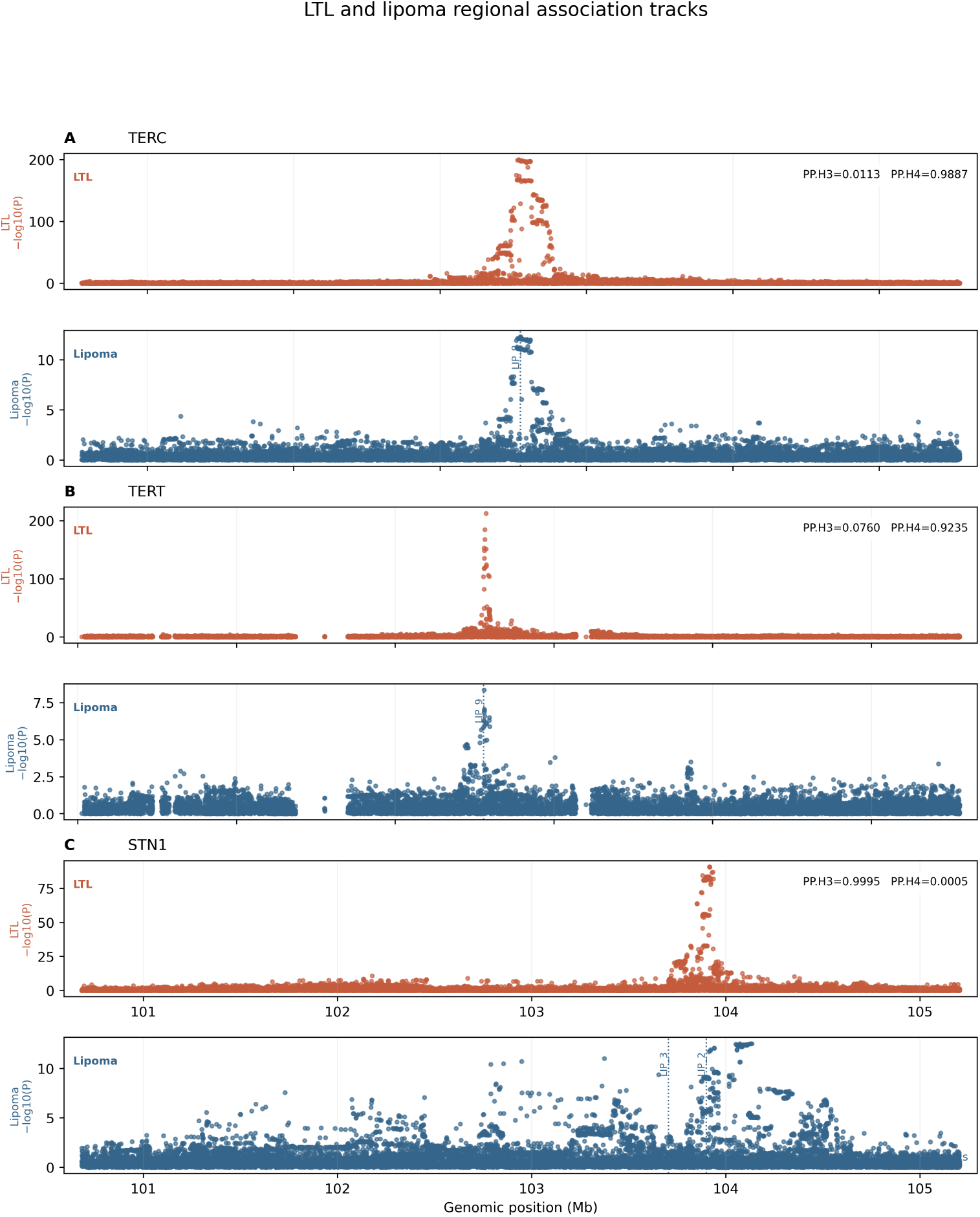
Regional association tracks for LTL and FinnGen lipoma susceptibility at TERC, TERT, and STN1/OBFC1. Each locus has vertically aligned upper LTL and lower lipoma tracks sharing genomic x coordinates but using independent y-axis scales. The x-axis is GRCh38 genomic position in Mb and each y-axis is −log10(P). Orange points are LTL and blue points are lipoma. Dotted blue vertical lines and labels identify FinnGen SuSiE signal leads; the STN1/OBFC1 region shows both chr10 lipoma signals. PP.H3 denotes distinct causal variants and PP.H4 denotes a shared causal variant under the coloc model. TERC: PP.H3=0.0113 and PP.H4=0.9887, supporting a shared regional component without unique shared-SNP or causal-gene resolution. TERT: PP.H3=0.0760 and PP.H4=0.9235, with discordant fine-mapping and moderate Finnish LD (r² approximately 0.515), leaving local architecture unresolved. STN1/OBFC1: PP.H3=0.9995 and PP.H4=0.00049, favoring distinct components in a two-signal lipoma region. No LD or PIP color gradient, credible-set shading, error bars, or causal- boundary encoding is plotted. LTL, leukocyte telomere length; PIP, posterior inclusion probability; LD, linkage disequilibrium; r², squared correlation; PP, posterior probability; SuSiE, Sum of Single Effects.

**Table 5.**
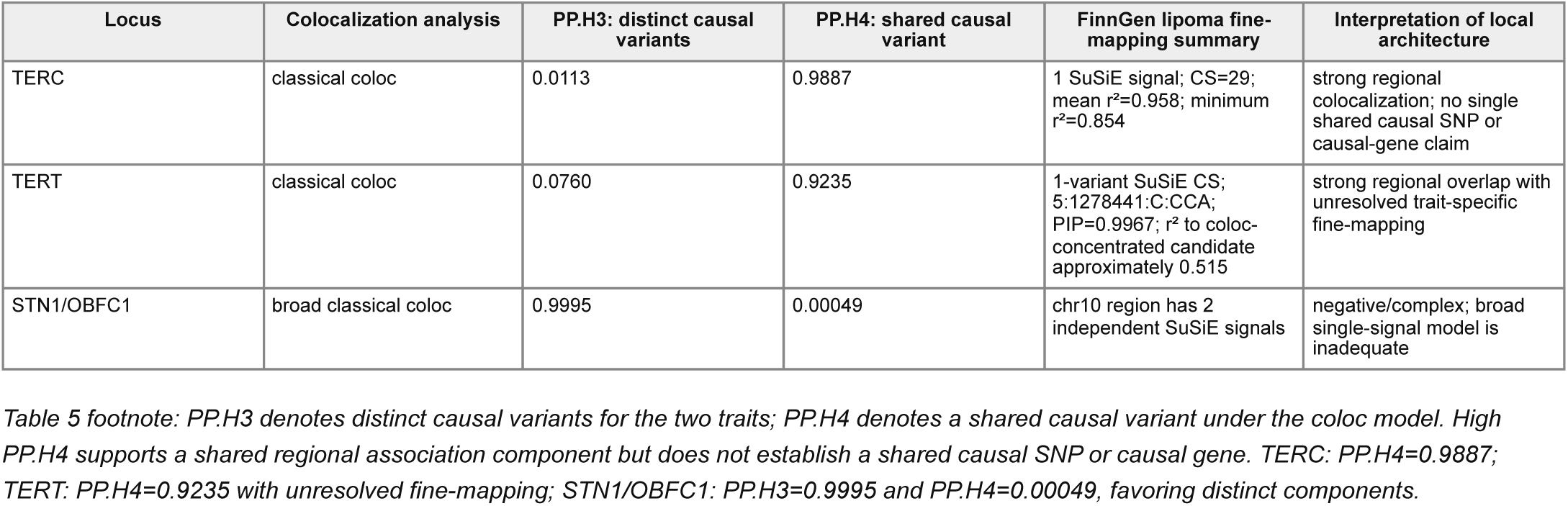
Colocalization and fine-mapping reveal heterogeneous local relationships between LTL and lipoma at TERC, TERT, and STN1.

footnote: PP.H3 denotes distinct causal variants for the two traits; PP.H4 denotes a shared causal variant under the coloc model.
| Locus | Colocalization analysis | PP.H3: distinct causal variants | PP.H4: shared causal variant | FinnGen lipoma fine-mapping summary | Interpretation of local architecture |
| --- | --- | --- | --- | --- | --- |
| TERC | classical coloc | 0.0113 | 0.9887 | 1 SuSiE signal; CS=29; mean $r^2=0.958$; minimum $r^2=0.854$ | strong regional colocalization; no single shared causal SNP or causal-gene claim |
| TERT | classical coloc | 0.0760 | 0.9235 | 1-variant SuSiE CS; 5:1278441:C:CCA; PIP=0.9967; $r^2$ to coloc-concentrated candidate approximately 0.515 | strong regional overlap with unresolved trait-specific fine-mapping |
| STN1/OBFC1 | broad classical coloc | 0.9995 | 0.00049 | chr10 region has 2 independent SuSiE signals | negative/complex; broad single-signal model is inadequate |

The strongest evidence for regional colocalization occurred at the *TERC* locus on chromosome 3. Classical colocalization strongly favored a shared association component (PP.H4=0.989) over distinct causal variants within the region (PP.H3=0.011). FinnGen fine-mapping independently identified a single high-purity lipoma SuSiE signal in the region, comprising 29 variants with average r²=0.958 and minimum r²=0.854. Together with the closely aligned regional LTL and lipoma association profiles, these findings provide strong evidence for a shared *TERC*-region association component. They do not, however, resolve a single shared causal variant or establish *TERC* itself as the causal lipoma gene.

The *TERT* region on chromosome 5 also showed strong regional overlap but less concordant fine-mapping. Classical colocalization favored a shared association (PP.H4=0.924; PP.H3=0.076), while FinnGen SuSiE resolved the lipoma association to a one-variant 95% credible set at 5:1278441:C:CCA (PIP=0.997). The variant carrying the greatest shared posterior probability in the colocalization analysis differed from the FinnGen lipoma fine-mapping candidate, and Finnish linkage disequilibrium between the key variants was only moderate (r2∼0.515). The *TERT* findings are therefore interpreted as strong regional overlap with unresolved trait-specific causal architecture rather than evidence for a single shared causal variant.

In contrast, the *STN1/OBFC1* region on chromosome 10 did not support a simple shared-signal model. Broad-region colocalization overwhelmingly favored distinct association components (PP.H3=0.9995; PP.H4=0.00049), while FinnGen fine-mapping identified two independent lipoma signals within the region. The current data therefore do not support a shared single association component between LTL and lipoma at *STN1/OBFC1*.

Collectively, these locus-level analyses reveal heterogeneous relationships between LTL and lipoma susceptibility. *TERC* provides the strongest local evidence for shared genetic architecture, *TERT* shows strong but incompletely resolved regional overlap, and *STN1/OBFC1* provides an important counterexample to a simple model in which every telomere-associated lipoma locus acts through the same shared genetic signal.

## Discussion

Allaire et al. identified lipoma as the strongest tumor association in their phenome-wide analysis of genetic predisposition toward longer telomeres, providing compelling initial evidence for a connection between inherited telomere biology and lipoma susceptibility [19]. The present study approaches that relationship from the complementary direction of lipoma genetics and provides several independent lines of convergent evidence. Lipoma susceptibility loci identified without reference to LTL genetics mapped to multiple genes involved in telomere maintenance and genome stability; genetically proxied longer LTL was strongly associated with increased lipoma susceptibility using All of Us exposure effects and replicated using independent UK Biobank effects; the association persisted after exclusion of variants assigned to canonical telomere/DDR genes and across anatomical sites; and locus-level analyses provided strong regional support at *TERC* while revealing more complex architectures at *TERT* and *STN1*. Taken together, the results place a common benign neoplasm within a broader biological trade-off between maintenance of cellular replicative capacity and protection from abnormal clonal growth. At the same time, substantial heterogeneity among MR estimates and individual loci argues against a single mechanism uniformly linking telomere length to lipoma formation.

### Telomere biology, aging, and benign neoplasia

Telomere attrition occupies an unusual position at the intersection of aging and tumor suppression. Progressive shortening restricts the number of divisions available to somatic cells and can arrest cells with oncogenic potential, but reduced proliferative capacity can also impair tissue maintenance and contribute to degenerative phenotypes of aging [23,32,33]. Campisi framed this broader relationship as antagonistic pleiotropy: cellular programs such as senescence can protect against cancer while contributing to age-related loss of regenerative function [32]. Human genetic studies provide a complementary population-level view. Haycock et al. found that genetically longer telomeres increased risk for several cancers while decreasing risk for several non-neoplastic diseases, explicitly highlighting a trade-off between cancer resistance and degenerative disease [23]. A recent synthesis by Stuart et al. further emphasizes normal telomere attrition as a broadly acting tumor-suppressive mechanism that limits cellular proliferation and tumor outgrowth [33].

Lipoma may be informative precisely because it lies outside the usual cancer-focused framing of this trade-off. A lipoma is a clonal neoplasm, but it is benign and has extremely limited malignant potential. The association between longer-LTL genetics and lipoma therefore suggests that the proliferative side of the telomere trade-off may influence the probability of benign clonal expansion as well as malignant transformation. In this view, the relevant continuum is not simply aging versus cancer, but preservation of cellular replicative competence versus increasingly stringent suppression of abnormal clonal growth.

The direction and distribution of the LTL association are consistent with the model proposed by Allaire et al., in which genetically longer telomeres increase cellular replicative potential and thereby extend the opportunity for tumor-promoting somatic events to occur before telomere-dependent senescence [19]. The lipoma results add a complementary dimension to that model because the independently derived germline architecture of lipoma susceptibility itself converges on telomere and genome-maintenance biology. Given the well-established diversity of somatic alterations in lipoma, an attractive hypothesis is that inherited replicative state modifies the probability that cells carrying any of several somatic alterations persist and undergo benign clonal expansion rather than specifying a particular somatic driver. Testing that model directly will require paired germline and tumor genomic data.

Historical observations of telomere dysfunction in adipocytic tumors are compatible with, but distinct from, this germline model. In 48 atypical lipomatous tumors, Mandahl et al. documented 12 clonal and 344 nonclonal telomeric associations with a nonrandom distribution across chromosome ends [5]. The authors considered whether chromosome-end instability might facilitate structural rearrangements while acknowledging that both phenomena could instead reflect broader genomic instability. Somatic telomere dysfunction should not be equated with inherited propensity toward longer LTL: tumor telomerase activity, tumor telomere length, chromosome-end instability, and genetically proxied leukocyte telomere length are related but distinct biological quantities. A cell with relatively high replicative competence could later acquire telomere dysfunction or other genomic instability during clonal evolution.

### Heterogeneous genetic routes connect telomere biology to lipoma susceptibility

The germline findings complement a longstanding literature establishing lipomas as genetically heterogeneous clonal neoplasms. Cytogenetic studies identified recurrent somatic routes involving chromosome 12q13–15, ring chromosomes, 6p, chromosome 13, and other abnormalities [1]. In the 188-tumor series, 55 sporadic lipomas had 12q13–15 abnormalities, 20 had supernumerary ring chromosomes, 11 had changes involving 6p or chromosome 13 without either of those features, and 14 had other aberrations; clonal evolution occurred in approximately 30% of tumors with relevant sampling information [1]. Subsequent work established *HMGA2* as an important target of 12q13–15 rearrangements [2–4], and modern sequencing has further demonstrated heterogeneity among ordinary lipomas rather than a single recurrent somatic driver [21]. Whereas that literature describes the somatic genetics of established tumors, the present results address a complementary question: what inherited biological state predisposes an individual to establish a benign adipocytic clone?

The distinction between inherited susceptibility and somatic driver may be especially relevant to multiple lipomas. In the same cytogenetic series, only one of 58 tumors from 18 patients with multiple lipomas had detectable karyotypic changes, leading Mandahl et al. to suggest a constitutional predisposition [1]. The population-level analyses presented here do not specifically address familial multiple lipomatosis and do not establish that common LTL-associated variants explain familial phenotypes. They do, however, support inherited variation in cellular maintenance and replicative biology as a component of lipoma susceptibility. Testing whether this architecture has larger effects in individuals with multiple tumors is an important future direction.

The locus-level analyses show why overlap between traits should not automatically be interpreted as a single pleiotropic effect. When two traits map to the same region, the pattern can arise because one causal variant affects both traits, because distinct causal variants are linked through LD, or because one or both traits have multiple causal signals. *TERC* provided the strongest local evidence consistent with a shared association component between LTL and lipoma, although the data do not identify a single shared causal variant. At *TERT*, high classical colocalization support contrasted with different fine-mapped candidates and only moderate LD between key variants, leaving linkage between distinct causal effects or more complex local architecture plausible. At *STN1*, the evidence strongly favored distinct association components. These contrasts show that convergence on telomere biology does not require every lipoma locus to act by pleiotropically altering LTL through the same causal variant.

Potential parallels with other benign mesenchymal tumors remain intriguing but unproven. Several lipoma-associated regions, including those near *TERC*, *TERT*, *STN1*, BET1-like Golgi vesicular membrane trafficking protein (*BET1L*), and ATM serine/threonine kinase (*ATM*), showed strong and directionally concordant associations with uterine leiomyoma. Such regional coincidence could reflect pleiotropy, linkage between distinct trait-specific variants, or chance overlap among association-rich regions. The systematic regional comparison did not show enrichment of lipoma–leiomyoma overlap, and native signal-level leiomyoma fine-mapping was unavailable for rigorous comparison. These observations therefore remain hypothesis-generating rather than evidence for a shared benign-mesenchymal genetic architecture.

### Limitations and future directions

Several limitations qualify the conclusions. Mendelian randomization relies on assumptions that cannot be fully demonstrated empirically. Variant-specific estimates were heterogeneous, and although weighted-median, MR-Egger, RadialMR, leave-one-out, directionality, and canonical-gene exclusion analyses supported the primary association, horizontal pleiotropy remains plausible. A nonsignificant MR-Egger intercept does not establish its absence. The MR results are therefore interpreted as showing that the genetic architecture underlying longer LTL is associated with lipoma susceptibility, not that experimentally increasing telomere length would cause lipoma.

The exposure data introduce additional constraints. All of Us and UK Biobank LTL effects are not documented on an identical standardized scale, so replication is strongest in the concordant variant effects and direction of association rather than equality of effect magnitudes. LTL is also a blood-derived phenotype, and genetic effects on leukocyte telomere length need not capture telomere regulation identically in adipose progenitor cells or other cells relevant to lipoma formation.

Several lipoma loci, particularly on chromosomes 20 and 22, remain poorly fine-mapped, and overlapping or nearby genes should not be treated as established causal genes. Functional annotation also lacked comprehensive regulatory, eQTL, and sQTL information and is best viewed as independent biological convergence rather than definitive gene assignment or pathway enrichment.

The most direct next step is to integrate inherited susceptibility with the somatic genomes of individual lipomas. Paired germline and tumor sequencing from individuals with solitary and multiple lipomas could test whether LTL-associated genetic backgrounds modify the probability, timing, or clonal expansion of particular somatic events. Larger lipoma GWASs, improved fine-mapping, and adipose-lineage regulatory data will also be needed to distinguish causal genes from nearby candidates and to determine whether the telomere-related signal acts through telomere length itself or through broader functions in DNA replication, repair, and cellular-state maintenance.

Together, these findings connect the independently derived genetic architecture of lipoma susceptibility with prior evidence for long-telomere tumor predisposition and with the broader trade-off between regenerative capacity and neoplastic restraint. Fine-mapped lipoma loci, replicated LTL genetic associations, anatomical generality, and heterogeneous locus-level relationships support a model in which inherited cellular replicative state may influence benign adipocytic clonal establishment or persistence without requiring a single telomere-dependent pathway. Lipoma therefore provides a potentially informative benign endpoint for understanding how the same biological systems that preserve proliferative capacity across the lifespan can also increase the opportunity for abnormal clones to expand.

## Data Availability

All derived data produced in this study, including data underlying the figures and tables, are publicly available at https://github.com/lowrylab/lipoma-telomere-genetics and are archived in the version 1.0.0 research compendium at Zenodo (https://doi.org/10.5281/zenodo.22651692). Large source GWAS datasets are not redistributed; their provenance and access information are documented in the repository source manifest. The All of Us and UK Biobank leukocyte telomere-length summary statistics used in this study are publicly available aggregate data. No individual-level participant data were accessed.

https://doi.org/10.5281/zenodo.22651692

https://doi.org/10.5281/zenodo.16689623

## Data and Code Availability

Analysis scripts, derived data underlying the figures and tables, detailed data-source provenance, reproducibility documentation, and the Supplementary Information are available at the public GitHub repository: https://github.com/lowrylab/lipoma-telomere-genetics. The research compendium accompanying this preprint is archived as version 1.0.0 at Zenodo: https://doi.org/10.5281/zenodo.22651692. Large or redistribution-restricted source GWAS datasets are not redistributed in the repository; their provenance and access information are documented in the repository source manifest. Public aggregate All of Us and UK Biobank LTL summary-statistics provenance is documented there. No individual-level All of Us participant data were accessed.

## Notes

### Competing Interest Statement

The authors have declared no competing interest.

### Author Declarations

This study used only publicly released aggregate genetic summary statistics and fine-mapping results that were available before initiation of the study. No individual-level participant data were accessed. FinnGen R13 GWAS and fine-mapping results were obtained from the FinnGen public data release: https://www.finngen.fi/en/access_results Release documentation: https://docs.finngen.fi/finngen-data-specifics/finngen-data-freezes-and-releases All of Us and UK Biobank leukocyte telomere-length GWAS summary statistics were obtained from the publicly released Nakao et al. Zenodo archive: https://doi.org/10.5281/zenodo.16689623 The analyses therefore used only aggregate, previously released research data. No individual-level All of Us, UK Biobank, or FinnGen participant data were accessed.

## References

1. Mandahl N, Höglund M, Mertens F, Rydholm A, Willén H, Brosjö O, Mitelman F. Cytogenetic aberrations in 188 benign and borderline adipose tissue tumors. Genes Chromosomes Cancer. 1994;9(3):207–215. DOI: 10.1002/gcc.2870090309. PMID: 7515663.

2. Mandahl N, Heim S, Johansson B, Bennet K, Mertens F, Olsson G, Rööser B, Rydholm A, Willén H, Mitelman F. Lipomas have characteristic structural chromosomal rearrangements of 12q13–q14. International Journal of Cancer. 1987;39(6):685–688. DOI: 10.1002/ijc.2910390605. PMID: 3473046.

3. Schoenmakers EFPM, Wanschura S, Mols R, Bullerdiek J, Van den Berghe H, Van de Ven WJ. Recurrent rearrangements in the high mobility group protein gene, HMGI-C, in benign mesenchymal tumours. Nature Genetics. 1995;10(4):436–444. DOI: 10.1038/ng0895-436. PMID: 7670494.

4. Fedele M, Battista S, Manfioletti G, Croce CM, Giancotti V, Fusco A. Role of the high mobility group A proteins in human lipomas. Carcinogenesis. 2001;22(10):1583–1591. DOI: 10.1093/carcin/22.10.1583. PMID: 11576996.

5. Mandahl N, Mertens F, Willén H, Rydholm A, Kreicbergs A, Mitelman F. Nonrandom pattern of telomeric associations in atypical lipomatous tumors with ring and giant marker chromosomes. Cancer Genetics and Cytogenetics. 1998;103(1):25–34. DOI: 10.1016/S0165-4608(97)00268-9. PMID: 9595041.

6. Codd V, et al. Identification of seven loci affecting mean telomere length and their association with disease. Nature Genetics. 2013;45(4):422–427. DOI: 10.1038/ng.2528. PMID: 23535734.

7. Nakao T, et al. Genomic, phenomic and geographic associations of leukocyte telomere length in the United States. Nature Genetics. 2026. DOI: 10.1038/s41588-026-02567-1. PMID: 41896353.

8. Bowden J, Davey Smith G, Burgess S. Mendelian randomization with invalid instruments: effect estimation and bias detection through Egger regression. International Journal of Epidemiology. 2015;44(2):512–525. DOI: 10.1093/ije/dyv080.

9. Bowden J, Davey Smith G, Haycock PC, Burgess S. Consistent estimation in Mendelian randomization with some invalid instruments using a weighted median estimator. Genetic Epidemiology. 2016;40(4):304–314. DOI: 10.1002/gepi.21965.

10. Verbanck M, Chen CY, Neale B, Do R. Detection of widespread horizontal pleiotropy in causal relationships inferred from Mendelian randomization. Nature Genetics. 2018;50:693–698. DOI: 10.1038/s41588-018-0099-7.

11. Hemani G, et al. The MR-Base platform supports systematic causal inference across the human phenome. eLife. 2018;7:e34408. DOI: 10.7554/eLife.34408.

12. Wang G, Sarkar A, Carbonetto P, Stephens M. A simple new approach to variable selection in regression, with application to genetic fine-mapping. JRSS Series B. 2020;82(5):1273–1300. DOI: 10.1111/rssb.12388.

13. Benner C, Spencer CCA, Havulinna AS, Salomaa V, Ripatti S, Pirinen M. FINEMAP: efficient variable selection using summary data from genome-wide association studies. Bioinformatics. 2016;32(10):1493–1501. DOI: 10.1093/bioinformatics/btw018.

14. Giambartolomei C, et al. Bayesian test for colocalisation between pairs of genetic association studies using summary statistics. PLoS Genetics. 2014;10(5):e1004383. DOI: 10.1371/journal.pgen.1004383.

15. Boccardi V, Razdan N, Kaplunov J, et al. Stn1 is critical for telomere maintenance and long-term viability of somatic human cells. Aging Cell. 2015;14(3):372–381. DOI: 10.1111/acel.12289. PMID: 25684230.

16. Bowden J, Spiller W, Del Greco M F, Sheehan N, Thompson J, Davey Smith G. Improving the visualization, interpretation and analysis of two-sample summary data Mendelian randomization via the Radial plot and Radial regression. International Journal of Epidemiology. 2018;47(4):1264–1278. DOI: 10.1093/ije/dyy101.

17. Uringa E-J, Lisaingo K, Pickett HA, Brind’Amour J, Rohde J-H, Zelensky A, Essers J, Lansdorp PM. RTEL1 contributes to DNA replication and repair and telomere maintenance. Molecular Biology of the Cell. 2012;23(14):2782– 2792. DOI: 10.1091/mbc.E12-03-0179. PMID: 22593209.

18. Klein JC, Mahapatra R, Hon GC, Wang RC. Identification of Associations with Dermatologic Diseases through a Focused GWAS of the UK Biobank. JID Innovations. 2025;5(1):100322. DOI: 10.1016/j.xjidi.2024.100322. PMID: 39624183.

19. Allaire P, Mayer J, Moat L, et al. Long-telomopathy is associated with tumor predisposition syndrome. medRxiv. 2024. DOI: 10.1101/2024.11.26.24318007. PMID: 39649603.

20. Marzyńska D, Żaba R, Lacka K. Lipomas: genetic basis of common skin lesions and their occurrence in rare diseases. Postępy Dermatologii i Alergologii. 2023;40(4):481–486. DOI: 10.5114/ada.2023.129529. PMID: 37692275.

21. Kanojia D, et al. Identification of somatic alterations in lipoma using whole exome sequencing. Scientific Reports. 2019;9:14416. DOI: 10.1038/s41598-019-50805-w. PMID: 31591430.

22. Mejia Granados CM, et al. Clinical and Molecular Investigation of Familial Multiple Lipomatosis: Variants in the HMGA2 Gene. Clinical, Cosmetic and Investigational Dermatology. 2020;13:47–54. DOI: 10.2147/CCID.S213139. PMID: 32021365.

23. Haycock PC, Burgess S, Nounu A, et al. Association Between Telomere Length and Risk of Cancer and Non-Neoplastic Diseases: A Mendelian Randomization Study. JAMA Oncology. 2017;3(5):636–651. DOI: 10.1001/jamaoncol.2016.5945. PMID: 28241208.

24. Zhang C, Doherty A, Burgess S, et al. Genetic determinants of telomere length and risk of common cancers: a Mendelian randomization study. Human Molecular Genetics. 2015;24(19):5356–5366. DOI: 10.1093/hmg/ddv252. PMID: 26138067.

25. Huang J, et al. Identifying Potential Causal Effects of Telomere Length on Health Outcomes: A Phenome-Wide Investigation and Mendelian Randomization Study. The Journals of Gerontology: Series A. 2024; 79(1):glad128. DOI: 10.1093/gerona/glad128. PMID: 37209418.

26. FinnGen. FinnGen Data Freezes and Releases; Data Releases 2025. FinnGen Handbook. R13 release documentation and production-library resource paths. Available at: https://docs.finngen.fi/finngen-data-specifics/finngen-data-freezes-and-releases and https://docs.finngen.fi/release-notes/data-releases-2025.

27. FINNGEN. Statistical fine-mapping pipeline in FinnGen. GitHub repository documentation. Available at: https://github.com/FINNGEN/finemapping-pipeline. Documents 3-Mb regions, LDstore2 in-sample dosage LD, FINEMAP and SuSiE, and maximum L=10 causal variants.

28. Hemani G, Tilling K, Davey Smith G. Orienting the causal relationship between imprecisely measured traits using GWAS summary data. PLoS Genetics. 2017;13(11):e1007081. DOI: 10.1371/journal.pgen.1007081. PMID: 29149188.

29. Yates A, Beal K, Keenan S, et al. The Ensembl REST API: Ensembl Data for Any Language. Bioinformatics. 2015;31(1):143–145. DOI: 10.1093/bioinformatics/btu613. PMID: 25236461.

30. Zerbino DR, Achuthan P, Akanni W, et al. Ensembl 2018. Nucleic Acids Research. 2018;46(D1):D754–D761. DOI: 10.1093/nar/gkx1098. PMID: 29155950.

31. Kolberg L, Raudvere U, Kuzmin I, et al. g:Profiler—interoperable web service for functional enrichment analysis and gene identifier mapping (2023 update). Nucleic Acids Research. 2023;51(W1):W207–W212. DOI: 10.1093/nar/gkad347.

32. Campisi J. Cancer and ageing: rival demons? Nature Reviews Cancer. 2003;3(5):339–349. DOI: 10.1038/nrc1073. PMID: 12724732.

33. Stuart A, Zhang B, Hockemeyer D, de Lange T. How telomere attrition protects against cancer. Nature Reviews Genetics. 2026. DOI: 10.1038/s41576-026-01001-w. PMID: 42680848.

