## Supplementary Information for "Genetic architecture of lipoma susceptibility implicates telomere biology"

### Supplementary Methods

#### Organization and data conventions

This Supplementary Information accompanies the V3 preprint candidate and provides the detailed results underlying the main text. It contains no additional analyses. Numerical values in the machine-readable tables are retained at their available precision; compact descriptions in this document use reader-facing rounding where appropriate.

All Mendelian-randomization (MR) estimates use the same 198 LTL-associated variants unless an item explicitly states otherwise. All of Us (AoU) is the primary exposure dataset and UK Biobank (UKB) is the independent exposure replication. AoU and UKB LTL effects are reported on their respective study-specific scales, not on a demonstrated common standardized-SD scale.

#### Variant-level MR and sensitivity procedures

The complete variant-level MR results retain the exposure and FinnGen lipoma outcome effect estimates, standard errors, alleles, allele frequencies, and harmonization fields used for the compact main Table 3. Separate tables provide the primary AoU analysis, the UKB exposure replication, single-variant estimates, and leave-one-out results.

Sensitivity results are separated by purpose. The canonical telomere/DDR analysis removes the prespecified 15 variants, leaving 183 variants. Leave-one-out results show the estimate after sequential omission of each contributing variant. RadialMR provides heterogeneity-oriented diagnostic and iterative estimates; flagged variants are diagnostics and were not automatically removed. MR-PRESSO outputs are reported as exploratory diagnostics only and were not used to select the primary LTL-associated variant set.

Heterogeneity, MR-Egger intercept, and Steiger directionality results are reported separately for AoU and UKB. Steiger directionality uses the binary-outcome approximation described in the main Methods and should be interpreted subject to that approximation.

#### Fine-mapping and locus-level analyses

The FinnGen lipoma fine-mapping supplement lists every variant in each of the nine independent SuSiE signals, together with signal identifiers, credible-set membership, posterior inclusion probabilities, and purity information. Locus-gene evidence is provided for direct-overlap and proximity-expanded mappings.

Regional association inputs for TERC, TERT, and STN1/OBFC1 contain the aligned LTL and lipoma summary statistics used for the displayed regional comparisons. PP.H3 represents distinct causal variants and PP.H4 represents a shared causal variant under the coloc model; neither posterior alone establishes a shared causal SNP or causal gene. The Finnish LD comparison supporting the TERT interpretation is reported with the locus-level outputs.

#### Functional annotation and enrichment

Functional annotation was performed using the prespecified direct-overlap and proximity-expanded mapping rules. The complete mapping tables retain variant, signal, gene, distance, and annotation fields. Enrichment outputs are provided separately for direct-overlap and proximity-expanded mappings. These results are descriptive evidence of biological convergence and do not constitute an independent genome-wide pathway test.

#### Exploratory leiomyoma comparison

The lipoma–leiomyoma materials provide the systematic regional comparisons, reciprocal cross-trait lookups, candidate follow-up loci, and architecture summaries generated from the available FinnGen summary statistics. The coarse regional permutation result is retained: 10 observed overlaps versus 13.2 expected, empirical P=1.0. Native leiomyoma fine-mapping and LD resources were not locally accessible, so no signal-level shared-causal interpretation is made.

### Supplementary Results

#### LTL-associated variant framework and variant-level MR

The published European LTL framework began with 210 variants; 198 were retained after harmonization to the exact AoU exposure data and FinnGen broad lipoma outcome. The corresponding UKB exposure replication uses the same 198 variants. Complete variant-level estimates, harmonization fields, and diagnostic outputs are available in Supplementary Tables S1–S5 and S7.

The canonical telomere/DDR sensitivity excludes exactly 15 variants and leaves 183. Leave-one-out results, RadialMR estimates and flags, and exploratory MR-PRESSO outputs are kept as distinct result classes so that diagnostic procedures are not confused with the definition of the primary LTL-associated variant set.

#### Lipoma genetic architecture and functional convergence

The nine independent FinnGen SuSiE signals span seven regions. Supplementary Table S2 provides the complete credible-set variants and locus-gene evidence. Supplementary Table S6 provides regional input data for the three loci emphasized in the main text. Supplementary functional outputs provide both mapping definitions and enrichment results supporting the qualitative convergence on telomere-maintenance and genome-maintenance biology.

#### Regional architecture at TERC, TERT, and STN1/OBFC1

The regional inputs and summary fields support the main interpretations: TERC shows strong regional colocalization without SNP-level causal resolution; TERT shows strong regional overlap with discordant trait-specific fine-mapping and moderate Finnish LD; and STN1/OBFC1 favors distinct association components in a complex two-signal region.

#### Exploratory lipoma–leiomyoma comparison

The exploratory comparison includes strong associations at several overlapping regions, but the regional-overlap permutation does not support enrichment. Because native leiomyoma signal-level fine-mapping and LD resources were unavailable locally, these results do not establish shared causal architecture.

### Supplementary Figures

#### Supplementary Figure S1. Human-supervised agentic research workflow and evidence-control process

This existing one-page schematic shows human scientific oversight, ChatGPT and Codex as research tools, iterative quality control, discrepancy resolution, and final evidence control. It does not assign independent scientific authority or authorship to either AI system. The figure is supplied in PDF and high-resolution PNG form in the repository.

No additional supplementary figures are included. A planned scatter/forest figure duplicates information already presented in main Figure 3 and was not separately produced. A planned leiomyoma figure was not available as an existing rendered output; no new figure was created.

### Supplementary Tables

#### Supplementary Table S1. Complete 198-variant LTL-associated variant manifest and harmonization quality-control fields

The table contains the retained 198 variants, genomic positions and alleles, exact AoU and UKB exposure effects, FinnGen lipoma outcome effects, allele frequencies, gene and telomere/DDR annotations, MR-PRESSO diagnostic status, and QC flags. It is the variant-level reference for the primary and replication MR analyses.

#### Supplementary Table S2. Complete FinnGen lipoma SuSiE credible-set variants and locus-gene evidence

Part A lists variants in the nine independent SuSiE signals, including signal identifiers, credible-set membership, PIP, and available purity fields. Part B lists direct-overlap and proximity-expanded gene-mapping evidence. PIP denotes posterior inclusion probability; SuSiE denotes Sum of Single Effects.

#### Supplementary Table S3. Complete MR estimates and variant-level diagnostics

This table family contains complete AoU and UKB MR results, single-variant estimates, and leave-one-out results. β is the MR estimate, SE is its standard error, CI is the confidence interval, and P is the P value. Effects are on the relevant study-specific LTL scale.

#### Supplementary Table S4. Canonical telomere/DDR exclusion sensitivity

This table identifies the exact 15 prespecified telomere/DNA-damage-response variants excluded in the sensitivity analysis and documents the resulting 183-variant analysis set. DDR denotes DNA-damage response.

#### Supplementary Table S5. Heterogeneity, MR-Egger, Steiger, RadialMR, and exploratory MR-PRESSO diagnostics

This table family contains Cochran Q and its P value, MR-Egger intercept results, Steiger directionality results, RadialMR iterative estimates and diagnostic flags, and MR-PRESSO outputs. Q is Cochran’s heterogeneity statistic. MR-PRESSO results are exploratory and were not used for primary variant selection.

#### Supplementary Table S6. Regional association inputs for TERC, TERT, and STN1/OBFC1

These tables contain aligned LTL and lipoma regional summary statistics and the fields used for the coloc comparisons. PP.H3 and PP.H4 are posterior probabilities for distinct and shared causal variants, respectively. A high PP.H4 supports a shared regional association component but does not establish a shared causal SNP or gene.

#### Supplementary Table S7. Exact AoU exposure-beta audit and IVW weighting decomposition

These tables compare published LTL-associated variant effects with exact AoU effects and document the decomposition of the earlier point-estimate discrepancy. They support the use of exact AoU summary-statistic effects in the primary analysis.

#### Supplementary Table S8. Exploratory lipoma–leiomyoma regional comparison

These tables contain the reciprocal cross-trait regional lookups, independent-signal summaries, architecture summaries, and follow-up candidates. The coarse permutation result is 10 observed overlaps versus 13.2 expected, empirical P=1.0. The comparison is exploratory and does not establish shared causal architecture.

#### Supplementary Table S9. Anatomical-site MR details

This table contains the complete anatomical-site MR summary for Limbs, Trunk, and Head/face/neck using the unchanged 198-variant framework. AoU and UKB estimates are identified separately and remain on study-specific LTL scales.

#### Machine-readable data map

The following links provide the complete machine-readable material corresponding to each supplementary item. They are provided as data resources rather than reproduced in full in this narrative document.

- S1: 198-variant LTL-associated variant manifest
- S2A: SuSiE credible-set variants and S2B: locus-gene evidence
- S3: All of Us MR outputs and UK Biobank MR outputs, including complete MR, single-variant, and leave-one-out results
- S4: canonical telomere/DDR exclusion set
- S5: MR diagnostics, including heterogeneity, MR-Egger, Steiger, RadialMR, and exploratory MR-PRESSO outputs
- S6: TERC, TERT, and STN1/OBFC1 regional inputs
- Functional gene mapping and direct-overlap enrichment / proximity-expanded enrichment
- S7: exact All of Us beta audit and IVW decomposition
- S8: exploratory lipoma–leiomyoma outputs and follow-up candidates
- S9: anatomical-site MR summary

### Data and code availability

The public repository provides the analysis scripts, exact software and resource metadata, data provenance, machine-readable tables, figure source tables, and the detailed AI/agentic workflow disclosure: https://github.com/lowrylab/lipoma-telomere-genetics. The research compendium accompanying this preprint is archived as version 1.0.0 at Zenodo. Restricted or large source GWAS files are not redistributed. Public aggregate All of Us and UKB summary-statistics provenance is documented in the repository source manifest. No individual-level All of Us participant data were accessed.
